# GWAS of stool frequency reveals genes, pathways, and cell types relevant to human gastrointestinal motility and irritable bowel syndrome

**DOI:** 10.1101/2020.06.17.20132555

**Authors:** Ferdinando Bonfiglio, Xingrong Liu, Christopher Smillie, Anita Pandit, Alexander Kurilshikov, Rodrigo Bacigalupe, Tenghao Zheng, Hieu Nim, Koldo Garcia-Etxebarria, Luis Bujanda, Anna Andreasson, Lars Agreus, Susanna Walter, Goncalo Abecasis, Chris Eijsbouts, Luke Jostins, Miles Parkes, David A Hughes, Nicholas Timpson, Jeroen Raes, Andre Franke, Nicholas A Kennedy, Aviv Regev, Alexandra Zhernakova, Magnus Simren, Michael Camilleri, Mauro D’Amato

**Affiliations:** School of Biological Sciences, Monash University, Clayton VIC, Australia; Unit of Clinical Epidemiology, Department of Medicine Solna, Karolinska Institutet, Stockholm, Sweden; Center for Molecular Medicine, Karolinska Institutet, Stockholm, Sweden; Klarman Cell Observatory, Broad Institute, Cambridge, MA, USA; Department of Biostatistics, University of Michigan, School of Public Health, Ann Arbor, MI, USA; Department of Genetics, University of Groningen, University Medical Center Groningen, Groningen, the Netherlands; Department of Microbiology and Immunology, Rega Instituut, KU Leuven, Leuven, Belgium; Center for Microbiology, VIB, Leuven, 3000, Belgium; Department of Gastrointestinal and Liver Diseases, Biodonostia HRI, San Sebastián, Spain; Centro de Investigación Biomédica en Red de Enfermedades Hepáticas y Digestivas (CIBERehd); Universidad del País Vasco (UPV/EHU), San Sebastian, Spain; Division of Clinical Medicine, Department of Medicine Solna, Karolinska Institutet, Stockholm, Sweden; Division of Family Medicine and Primary Care, Department of Neurobiology, Care Sciences and Society, Karolinska Institutet, Stockholm, Sweden; Division of Neuro and Inflammation Science, Department of Clinical and Experimental Medicine, Linköping University, Linköping, Sweden; Wellcome Centre for Human Genetics, Nuffield Department of Medicine, University of Oxford, Oxford, UK; Big Data Institute, Li Ka Shing Centre for Health Information and Discovery, University of Oxford, Oxford, UK; Kennedy Institute of Rheumatology, University of Oxford, Oxford, UK; Christ Church, University of Oxford, Oxford, UK; Division of Gastroenterology, Department of Medicine, University of Cambridge, UK; MRC Integrative Epidemiology Unit at University of Bristol, Bristol, UK; Population Health Sciences, Bristol Medical School, University of Bristol, Bristol, UK; Institute of Clinical Molecular Biology, Christian-Albrechts-University of Kiel, Kiel, Germany; IBD Pharmacogenetics, College of Medicine and Health, University of Exeter, Exeter, UK; Dept of Internal Medicine & Clinical Nutrition, Institute of Medicine, Sahlgrenska Academy, University of Gothenburg, Gothenburg, Sweden; Clinical Enteric Neuroscience Translational and Epidemiological Research (CENTER), and Division of Gastroenterology and Hepatology, Department of Medicine, Mayo Clinic, Rochester, MN, USA; IKERBASQUE, Basque Foundation for Science, Bilbao, Spain

**Keywords:** Intestinal motility, genetics, irritable bowel syndrome

## Abstract

**Objective:** Gut dysmotility is associated with constipation, diarrhea, and functional gastrointestinal disorders (FGID) like irritable bowel syndrome (IBS). Its molecular underpinnings, and their anomalies in FGID disorders are poorly characterized, hence we sought to gain mechanistic insight through a large-scale genetic investigation.

**Design:** We used stool frequency (STL-FRQ) as a (surrogate) quantitative trait to study the genetics of gut motility, exploiting questionnaire and genotype data from UK Biobank and four smaller population-based cohorts (LifeLines-Deep, Genes for Good, Flemish Gut Flora Project and PopCol), in a GWAS meta-analysis spanning 8,817,117 high-quality SNP markers and 167,875 individuals of European descent.

**Results:** We identify 13 genome-wide significant loci (P≤5.0×10^−8^) harboring prioritized genes that are: i) involved in sensory perception and neurotransmitter/neuropeptide signaling; ii) enriched for their expression in enteric motor neurons associated with the control of peristalsis (P=7.0×10^−8^) iii) previously linked to other traits and conditions, including GI motility and dysmotility syndromes, and the response to their pharmacological treatment. The genetic architecture of STL-FRQ most strongly correlates with that of IBS (r_g_=0.42; P=1.1×10^−3^). In UK Biobank, the risk of IBS with diarrhea was 4x higher in individuals from the top 1% of the distribution of polygenic scores (PGS) computed based on STL-FRQ GWAS summary statistics (ORs=4.14; P=1.2×10^−97^).

**Conclusion:** We identify loci harboring genes with a plausible role in GI motility, possibly acting via neurotransmission and similar pathways in specialized enteric neurons. The demonstrated relevance of these findings to IBS warrants further study for the identification of actionable pathomechanisms in the dysmotility syndromes.

## INTRODUCTION

Gastrointestinal (GI) motility is essential to digestion, nutrients absorption and overall human health, including bi-directional host-microbiome interactions.[1,2] Gut dysmotility and altered peristalsis are observed in constipation, diarrhea, and common functional GI disorders (FGIDs) like irritable bowel syndrome (IBS), which affect a large portion of the population and pose a remarkable socio-economic and healthcare burden.[3,4] While showing considerable symptoms overlap, most FGIDs are associated with some degree of GI motor dysfunction, possibly best exemplified by the observation that colonic transit time is generally delayed in patients with constipation-predominant IBS (IBS-C), and accelerated in patients with diarrhea-predominant IBS (IBS-D).[5,6] Dysmotility plausibly represents a key pathogenetic mechanism contributing to several GI conditions across a continuum ranging from mild symptoms, through functional disorders, and extreme cases of severely impaired peristalsis as observed in chronic idiopathic intestinal pseudo-obstruction.[7] There is only incomplete understanding of the physiological mechanisms regulating intestinal motility, and their perturbation in the dysmotility syndromes. While some medications (opioids, antidepressants, laxative and others) are known to influence GI motility, intrinsic triggers are generally believed to come from disturbed communication along the gut-brain axis, inflammatory or degenerative processes, and overstimulation of visceral sensory pathways that ultimately affect the gut musculature and local GI motor function via the enteric nervous system (ENS).[8] Therapeutic options are limited in the dysmotility syndromes, and rely on targeting specific symptoms rather than (currently unknown) underlying mechanisms. Genetic research may reveal biological pathways amenable to therapeutic exploitation, and some evidence of heritability can be derived for gut motility from previous studies in relation to colonic transit time measured with detectable tracers.[9–11] These studies, however, were largely underpowered to capture reliable genetic effects, as they lacked replication and focused on few DNA variants from individual candidate genes in small cohorts of IBS patients, hence results are not conclusive or transferable to the general population. As of today, no large-scale study has been performed to identify the genetic determinants of gut motility.

Direct assessment of GI motility in humans requires demanding clinical procedures (like transit time scintigraphy or the radiopaque markers method), which are exclusively performed to support patient diagnosis and therapeutic management, and are therefore not suitable for large-scale population-wide genetic surveys.[12] However, stool consistency and, to a lesser extent, stool frequency (STL-FRQ which refers to the number of bowel movements over a period of time) are valuable indicators of bowel function that correlate with colonic transit time, and can be practically recorded based on questionnaire- or diary survey-based approaches.[13,14] These represent therefore practical surrogate tools that can be adopted and scaled for studying GI motility at the population level thanks to their ease of data collection. Precedent for this approach comes, for instance, from a similar strategy recently applied in a genome-wide association study (GWAS) of IBS as self-reported condition from questionnaire data.[15]

Here, we leverage data from UK Biobank and four smaller population-based cohorts (LifeLines-Deep, Genes for Good, Flemish Gut Flora Project and PopCol) for a STL-FRQ GWAS meta-analysis across 8,817,117 high-quality single nucleotide polymorphism (SNP) markers in a total of 167,875 individuals of European descent. We identify 13 loci that harbor genes associated with pathways and cell types plausibly involved in the control of GI motility in humans, and provide compelling evidence of the relevance of these findings to IBS. The identification of genetic factors predisposing to altered gut motility may allow early identification of individuals at higher risk of FGID and, more importantly, therapeutically actionable pathways that may be targeted for the delineation of alternative treatment options.

## MATERIALS AND METHODS

### STUDY COHORTS

We studied phenotype and genotype data in 460,734 individuals from 5 population-based cohorts: UK Biobank (UKBB), LifeLines-DEEP cohort (LLD), The Genes for Good study (GFG), the Flemish Gut Flora Project (FGFP) and the Population-based Colonoscopy (PopCol) study. Health-related information was derived from questionnaires and participants’ electronic medical records, with STL-FRQ defined as the number of stool passes per day, after data harmonization. Selected genotype data was also studied in relation to colonic transit time (CTT) in a small cohort of 160 IBS patients from Sweden. A detailed description of all cohorts is reported in Supplementary Table 1 and the Supplementary Methods.

### GWAS META-ANALYSIS

A common GWAS pipeline was applied to individual cohorts based on mixed linear models and high quality (INFO>0.8) common (MAF>0.01) markers. A fixed-effect meta-analysis based on the inverse-variance weighted method was performed on a total of 167,875 individuals and 8,817,117 markers. In the analysis of CTT, STL-FRQ GWAS effect alleles were tested for association with linear regression adjusting for age, sex and first 10 principal components. A summary of relevant data, quality control measures, and analytical procedures is reported in the Supplementary Methods.

## FUNCTIONAL ANNOTATION OF STL-FRQ LOCI

### Locus definition and content

Annotation of *STL-FRQ* loci was done with FUMA v1.3.5 (https://fuma.ctglab.nl/), based on GWAS meta-analysis summary statistics. Independent association signals were identified based on SNP P-value (≤5.0×10^−8^) and linkage disequilibrium (LD) between markers (r2<0.4). Association signals were merged into a single locus for LD blocks closer than 250kb apart. Gene content at STL-FRQ loci was annotated based on positional and expression quantitative trait loci (eQTL) mapping, also with FUMA using default parameters and false discovery rate (FDR) P<0.05.

### Fine mapping

Fine-mapping was performed for the 13 genome-wide significant loci using FINEMAP v1.3,[16] with z-scores from the STL-FRQ GWAS meta-analysis and LD matrices derived from the genotype probabilities (.bgen files) of UK Biobank data. Specific eQTL traits associated with fine mapped SNPs were identified based on data from GTEx v8.[17]

## ENRICHMENT ANALYSES

### Gene-set and pathway enrichment analyses

Functional enrichment of STL-FRQ associated genes (as from positional and eQTL mapping with FUMA) was evaluated using GeneNetwork v2.0 (www.genenetwork.nl), in relation to KEGG pathways and Gene Ontology (GO) terms, using pre-computed co-regulation Z-scores and a Mann-Whitney U-test with FDR correction. Enrichment of molecular pathways from the REACTOME libraries was tested using PASCAL with STL-FRQ GWAS meta-analysis summary statistics, and default parameters including type I error control.[18]

### Cell-type enrichment analyses

Look up of STL-FRQ gene expression was done on previously reported single cell RNA-seq (scRNA-seq) and Ribosomes And Intact SIngle Nucleus isolation (RAISIN) RNA-seq data from human colonic mucosa and muscularis propria,[19,20] in relation to 76 cell types including immune, stromal and enteric neurons, among others. As described,[20] enteric neurons were partitioned into 5 classes based on the expression of major neurotransmitters/neuropeptides (*CHAT, SLC5A7, NOS1, VIP*) and other known markers: putative sensory neurons (PSN), interneurons (PIN subsets 1 and 2), secretomotor/vasodilator neurons (PSVN), and excitatory (PEMN subsets 1 and 2) and inhibitory motor neurons (PIMN subsets 1-5). Enrichment tests, comparing the expression of STL-FRQ genes versus background genes in enteric and motor neurons, were conducted using Fisher’s Exact test, controlling for type 1 error by FDR adjustment.

## CROSS-TRAIT ANALYSES

### Lookup of STL-FRQ GWAS signals in other traits

The GWAS catalog[21] and PhenoScanner[22] were screened with STL-FRQ lead SNPs and their high LD proxies (r2>0.8) in order to highlight associations (P≤5.0×10^−8^) with other traits. Associations were plotted with Circlize (cran.r-project.org/web/packages/circlize).

### Genetic correlations

STL-FRQ SNP heritability (h^2^ _SNP_) and genetic correlation (r_g_) between STL-FRQ and other complex traits were estimated using LD score regression (LDSC v1.0.1),[23] implemented in the CTG-VL platform (vl.genoma.io), which integrates public summary statistics of 1,387 traits from multiple repositories. Tests for statistical significance were FDR adjusted to control for type I errors.

## POLYGENIC SCORE ANALYSES

Polygenic scores (PGS) based on a pruning and thresholding approach were built using PRSice-2.[24] Effect estimates and corresponding standard errors from the STL-FRQ GWAS meta-analysis were used as the base dataset to generate weights, and then applied to IBS traits from UK Biobank (including subtypes) to derive PGS using PRSice-2 default settings. To account for the differences in the numbers of variants per cohort, a normalized polygenic score (mean=0, SD=1) was created per cohort. Student’s t-test was employed to determine the significance of the difference between the mean PGSs in IBS and controls. PGSs were binned into percentiles and the subset of IBS patients within a given magnitude of increased STL-FRQ PGS (top percentiles) was compared to the reminder of the population in a logistic regression adjusting for sex, age, the first 10 PCs and genotyping array.

## RESULTS

### STL-FRQ GWAS meta-analysis

The distribution of STL-FRQ (harmonized to the number of stool passes per day) was similar in the studied cohorts, with average ranging from 1.12 in GFG to 1.42 in UKBB and PopCol. Independent GWAS were carried out in individual cohorts using a common pipeline (Supplementary Methods), and later included in a meta-analysis encompassing 167,875 participants and 8,817,117 high-quality SNP markers. The STL-FRQ GWAS meta-analysis showed no population stratification (LDSC intercept = 1.02, see Methods), and identified 3751 genome-wide-significant associations (P≤5.0×10^−8^) from 13 independent loci (Table 1, Figure 1 and Supplementary Figure S1). The strongest signal was detected for marker rs12273363 on chromosome 11 (P=4.8×10^−21^), in proximity of the BDNF gene.

**Table 1.** STL-FRQ GWAS meta-analysis and fine mapping results

| Chr | Lead SNP | Start-end (bp) | EA | OA | EAF | Beta (SE) # | P | Nearest gene (other genes) * | Most likely causal SNP (% probability) ^ |
| --- | --- | --- | --- | --- | --- | --- | --- | --- | --- |
| 1 | rs11240503 | 205469956-205485290 | A | G | 0.300 | 0.018 (0.003) | 7.8E-09 | CDK18 (5) | <u>rs11240503</u> (0.588) |
| 5 | rs39819 | 122032544-122636855 | A | G | 0.671 | 0.018 (0.003) | 1.2E-09 | SNX24 (3) | - |
| 5 | rs13162291 | 154119471-154448827 | A | G | 0.191 | 0.020 (0.004) | 2.7E-08 | MRPL22 (5) | <u>rs13162291</u> (0.835) |
| 7 | rs12700026 | 2554037-2605424 | A | C | 0.890 | -0.029 (0.005) | 1.4E-10 | LFNG (5) | rs12700026, rs12700027 (0.350) |
| 7 | rs4556017 | 99919517-100678086 | T | C | 0.853 | 0.024 (0.004) | 1.0E-09 | MUC12 (46) | <u>rs4556017</u> (0.951) |
| 8 | rs10957534 | 71482998-72012331 | C | G | 0.367 | -0.016 (0.003) | 1.3E-08 | (5) | - |
| 11 | rs6486216 | 14980848-15127148 | T | C | 0.276 | 0.018 (0.003) | 1.1E-08 | CALCB (5) | - |
| 11 | rs12273363 | 27455582-27748493 | T | C | 0.795 | 0.032 (0.003) | 4.8E-21 | BDNF (3) | <u>rs12273363</u> (0.525) |
| 12 | rs11176001 | 66317487-66410673 | A | C | 0.132 | 0.034 (0.004) | 1.6E-16 | HMGA2 (1) | rs11176001 (0.392) |
| 12 | rs10492268 | 98298807-98506148 | T | C | 0.552 | 0.016 (0.003) | 1.6E-08 | (1) | rs10492268 (0.187) |
| 12 | rs3858648 | 115861753-115950227 | A | C | 0.508 | -0.016 (0.003) | 1.2E-08 |  | rs3858648 (0.077) |
| 17 | rs2732706 | 43460181-44865603 | T | C | 0.221 | 0.024 (0.003) | 4.4E-12 | ARL17A (109) | - |
| 22 | rs5757162 | 38869463-39152412 | T | C | 0.286 | 0.017 (0.003) | 4.0E-08 | FAM227A (21) | - |

Chr: chromosome, EA: effect allele; OA: other allele; EAF: effect allele frequency

**Figure 1.**
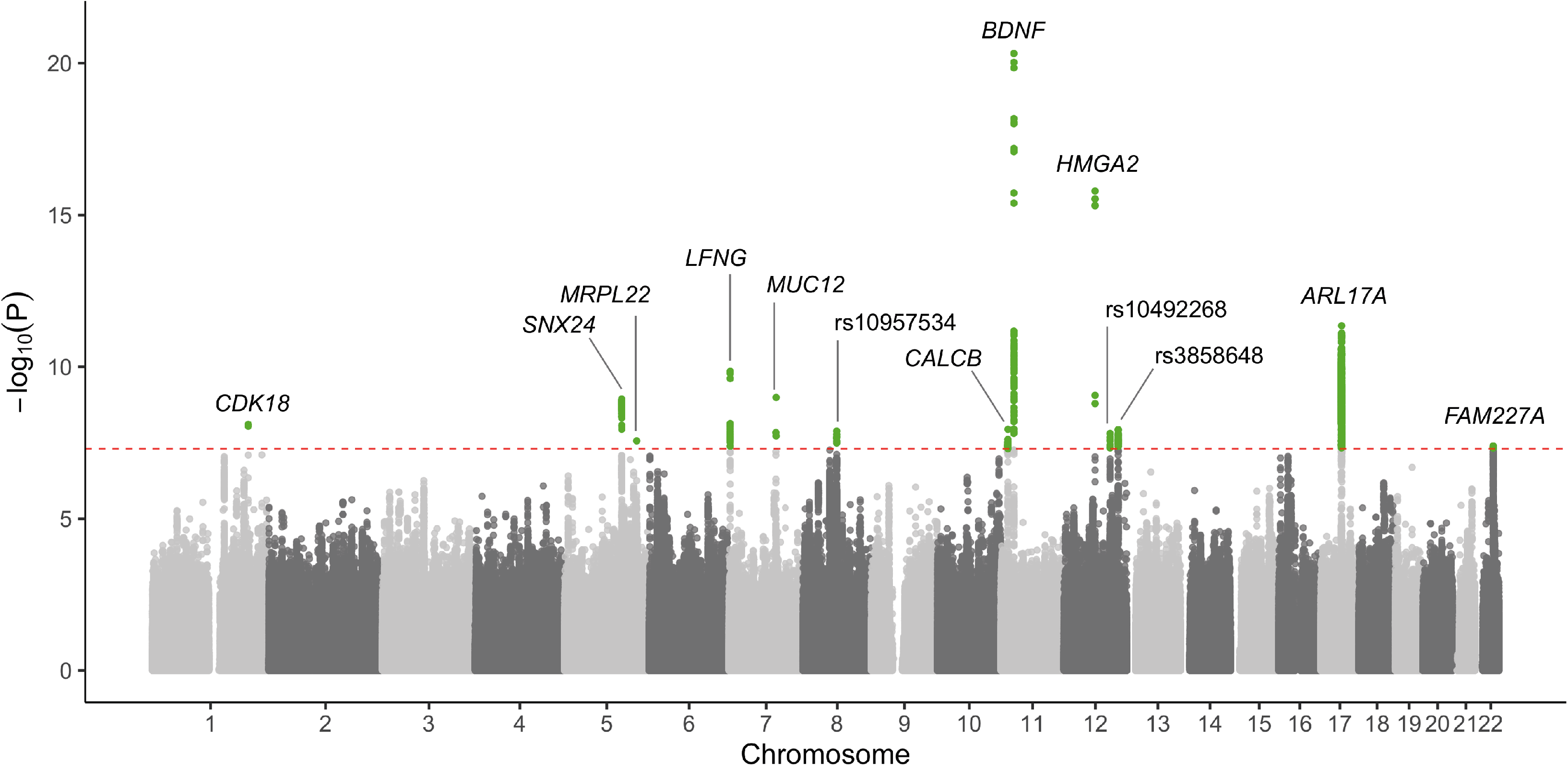
Manhattan plot of STL-FRQ GWAS meta-analysis results. GWAS association signals (−log_10_ P) are reported for SNP markers across all chromosomes shown in alternate gray colors. Significance level corresponding to genome wide (P=5.0×10^−8^) threshold is indicated with a dashed red horizontal line. For each independent association signal, the nearest gene (within 100kb, otherwise the lead SNP) is reported. Genome-wide significant markers are highlighted in green.

### Gene-set and pathway enrichment analyses

In order to obtain biological insight from the observed associations, we analysed STL-FRQ GWAS data with a computational pipeline for the functional annotation of associated loci. FUMA was used to define STL-FRQ GWAS loci, their boundaries and respective gene content, based on positional and eQTL mapping (see Methods). Several relevant genes were located at the associated loci (Supplementary Table 2), therefore we proceeded to perform gene-set and pathway enrichment analysis. GeneNetwork analysis revealed significant (FDR P<0.01) enrichment for relevant KEGG pathways including “*neuroactive ligand receptor interaction”*, and GO terms “*detection of chemical stimulus involved in sensory perception” and* “*neuropeptide signaling pathway”* (Figure 2 and Supplementary Table 3). Similarly, PASCAL pathway-level analysis highlighted “*neurotransmitter receptor binding and downstream transmission in the postsynaptic cell”* and “s*erotonin receptors”* as the top enriched REACTOME pathways (Supplementary Table 4).

**Figure 2.**
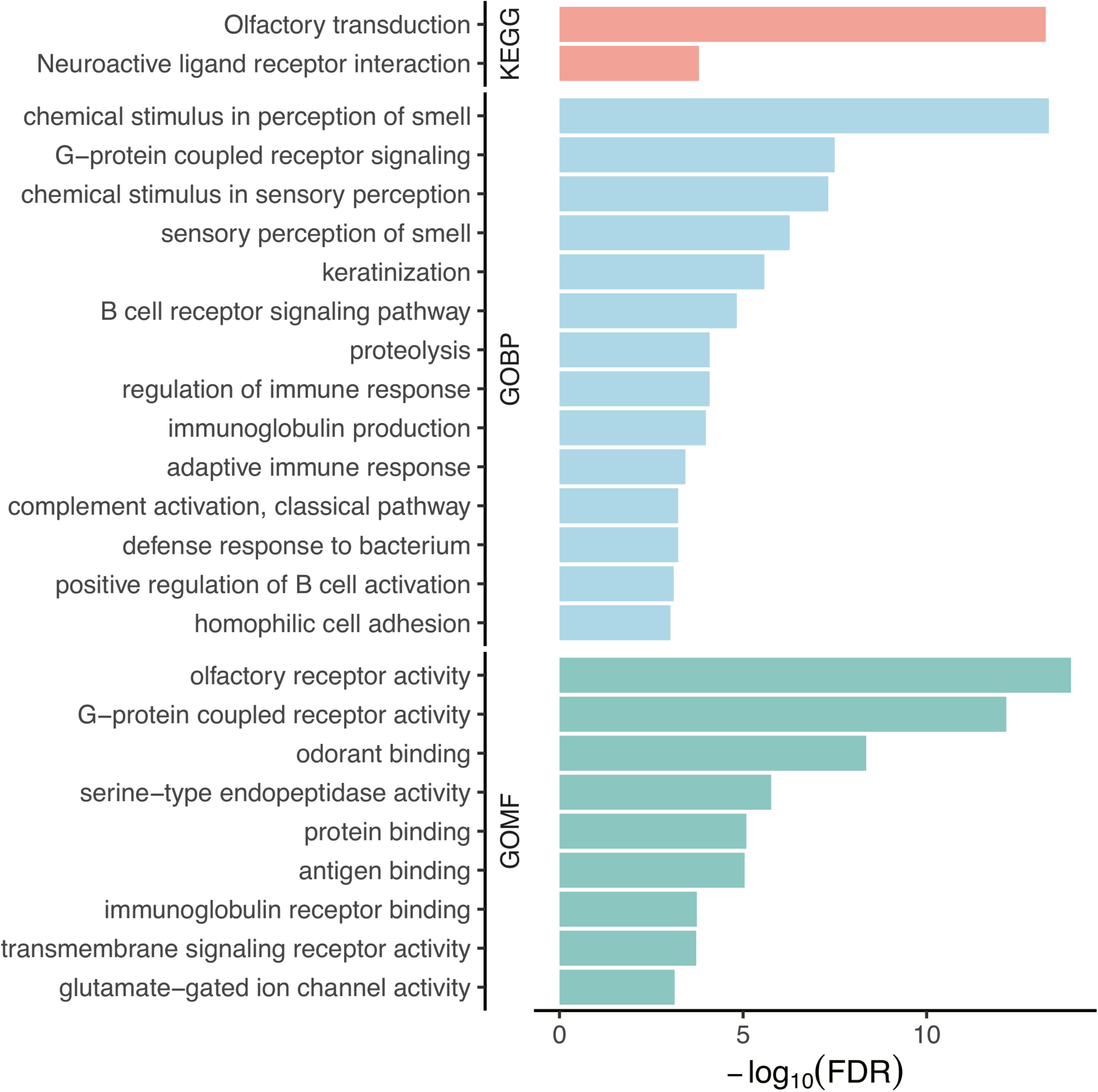
Gene set enrichment analysis results. Top significant findings from GeneNetwork analysis ranked by -log_10_ FDR adjusted P and color-coded according to KEGG pathways, Gene Ontology biological process (GOBP) and molecular function (GOMF) categories.

### Functional annotation at the single cell level

STL-FRQ genes did not show any preferential tissue expression based on FUMA or DEPICT analyses (not shown), hence we turned to study cell-type specific expression using single cell transcriptomic data available from human colonic mucosa and muscularis propria.[19,20] Look up of STL-FRQ gene expression was carried out using scRNA-seq and RAISINs RNA-seq data (see Methods), in relation to immune, epithelial, stromal and glial cells, muscle cells, and 11 subtypes of enteric neurons grouped into 5 major classes based on the relative expression of major neurotransmitters neuropeptides and other markers (Figure 3 and Supplementary Figure S2). Of note, the expression of STL-FRQ associated genes was strongly enriched in enteric neurons (FDR P=1.4×10^−3^), and more so in specific putative excitatory and inhibitory motor neurons (PEMN and PIMN subtypes, FDR P=7.0×10^− 8^) reportedly involved in the control of peristalsis.[20] Hence, functional annotation of STL-FRQ genes at the single cell level points to a potential role in controlling human gut motility through the involvement of specialized neuronal populations.

**Figure 3.**
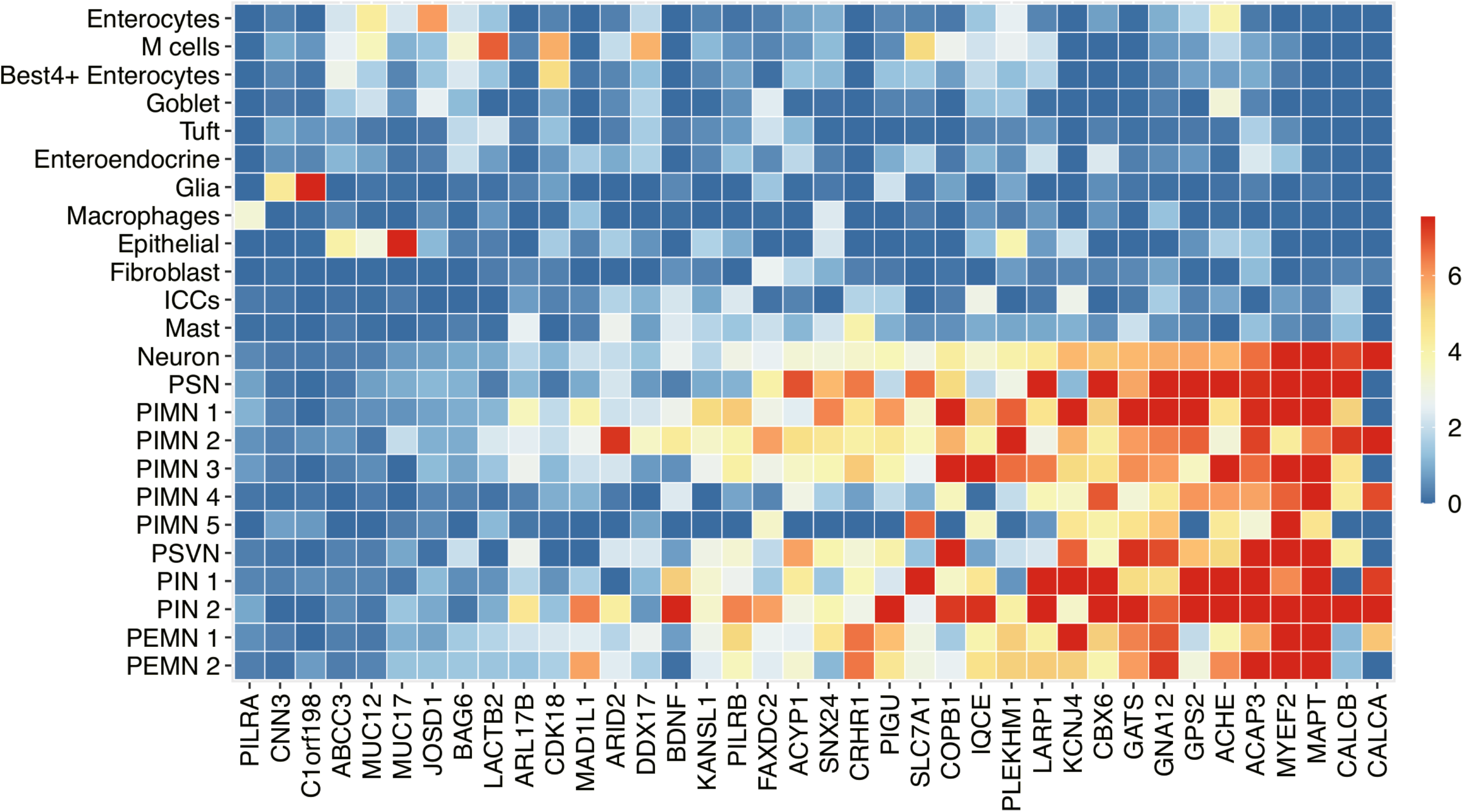
Heatmap of *STL-FRQ* gene expression in colonic cells. A selection of *STL-FRQ* genes is reported for their expression in relevant cell types from colonic mucosa and colonic muscularis, ordered according to increasing expression. The heatmap displays log2(TP10K+1) transformed data, and the expression of each gene is scaled across all cells and shown in color scale ranging from 0 to the 99th data quantile (to avoid high/low expressed genes dominating the heatmap). ICCs: interstitial cells of Cajal; PSN: putative sensory neurons; PEMN: putative excitatory motor neurons; PIMN: putative inhibitory motor neurons; PIN: putative interneurons; PSVN: secretomotor/vasodilator neurons. Cell types, neuron types and subtypes are classified as defined previously in Drokhlyansky et al. (Ref [20]).

### Prioritization of causative genes

FINEMAP analysis of candidate causative SNPs from STL-FRQ loci mapped 4/13 signals at single-marker resolution with >50% probability (Table 1). Variants rs4556017 and rs13162291 were mapped with highest confidence (respective probabilities 95.1% and 83.5%) and are both associated with eQTLs in multiple tissues (Supplementary Figure S3). In particular, rs4556017 shows eQTLs for the acetylcholinesterase *ACHE*, and rs13162291 eQTLs for the fatty acid hydroxylase *FAXDC2*, two genes expressed in enteric and motor neurons (Figure 3). The SNP rs11240503 is associated with a colon-specific eQTL for *CDK18* (Supplementary Figure S3), a protein kinase expressed in colonic M cells and BEST4+ enterocytes (Figure 3). Finally, the rs12273363 marker is associated with eQTLs for a long non coding antisense RNA (*BDNF-AS*, Supplementary Figure S3) modulating the expression of the brain-derived neurotrophic factor BDNF.[25] Of note, rs12273363 emerged as top GWAS signal in our meta-analysis (P=4.8×10^−21^, Table 1), and was also associated with consistent genetic effects on CTT measured by the radiopaque method in a small set of 160 IBS individuals (P=0.036, with the T allele associated both with more frequent stools and faster transit; Table 1 and Supplementary Table 5). Although fine mapping did not highlight most likely causative variants at other loci, their gene content includes candidate genes of known relevance to GI motility and dysmotility syndromes, like neuropeptides/neurotransmitters and their receptors (*CALCA*/*CALCB, CRHR1*), ion channels (*KCNJ4*), tight junction proteins (*CLDN15*) and others (Supplementary Table 2). Genetic variation at these loci also appears to affect gene expression across several tissues, as evidenced by eQTL analysis (Supplementary Figure S4).

### Correlations with other disease and traits

A lookup of STL-FRQ association signals in publicly available GWAS data suggested some of the 13 loci to be relevant to other traits and diseases across multiple domains, including health outcomes, lifestyle factors and anthropometric traits (Figure 4A). In particular, the loci tagged by markers rs12273363 and rs2732706 showed the largest number of associations, mostly with anthropometric and psychiatric traits, respectively (Supplementary Table 6).

**Figure 4.**
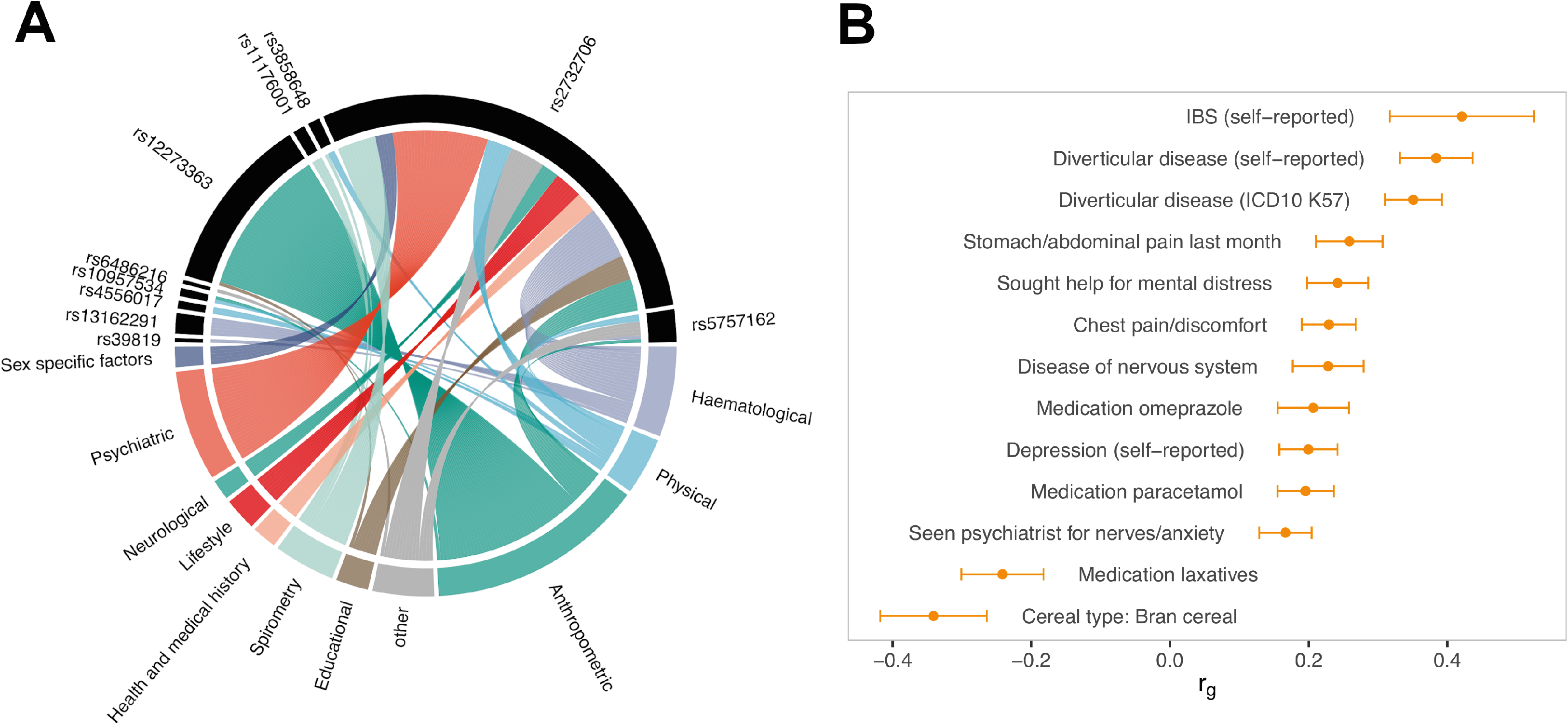
Cross-trait analysis of STL-FRQ GWAS results. A) Circus plot showing previously reported genome-wide significant associations (P=5.0×10^−8^) at the 13 STL-FRQ loci (lead SNPs or r2>0.8 LD proxies, see Methods). Associations are grouped by category, and the ribbon size is proportional to the number of associated traits in that category. In order to avoid redundancy, for multiple markers (lead or proxy SNP) linked to the same trait only the one with the lowest P is reported. No association were detected for 3 loci. B) Results obtained in the LDSC analyses of genetic correlation, in relation to a selection of most relevant traits (full results reported in Supplementary Table 7).

Evidence of genetic correlation with other conditions and traits was obtained from broader analyses of STL-FRQ GWAS summary statistics using LDSC analysis, which estimated SNP-based STL-FRQ heritability around 7% (h^2^_SNP_=0.073). When screening publicly available GWAS data (see Methods), strongest correlation was observed for IBS (r_g_=0.42, FDR P=5.1×10^−5^), while additional significant findings were obtained for other gastrointestinal (diverticular disease, use of proton pump inhibitors - Omeprazole) and psychiatric (anxiety, depression) traits, as well as a number of traits associated with pain and fatigue (Figure 4B and Supplementary Table 7), comorbidities often seen in dysmotility syndromes like IBS. Typifying inverse correlations were also detected for dietary fibers (bran cereals) and laxatives (dulcolax), which are usually consumed to avoid or relieve constipation, indeed a trait at or near the root of the STL-FRQ distribution tested here.

### STL-FRQ polygenic scores and irritable bowel syndrome

We further explored the relevance of STL-FRQ GWAS findings to IBS by computing PGS with PRSice-2 (see Methods) using STL-FRQ GWAS summary statistics. We studied STL-FRQ PGS in relation to IBS and its subtypes (constipation, IBS-C; diarrhea, IBS-D and mixed, IBS-M) defined according to gold-standard consensus Rome III criteria,[26] using data available for a subset of 164,979 UK Biobank participants who filled a digestive health questionnaire (the same used for the derivation of STL-FRQ in UK Biobank, Supplementary Methods). PGS distribution was significantly different in IBS vs asymptomatic individuals, most pronouncedly for IBS-D (mean PGS 0.456 vs -0.022 in cases and controls; P<1×10^−300^) (Figure 5).

**Figure 5.**
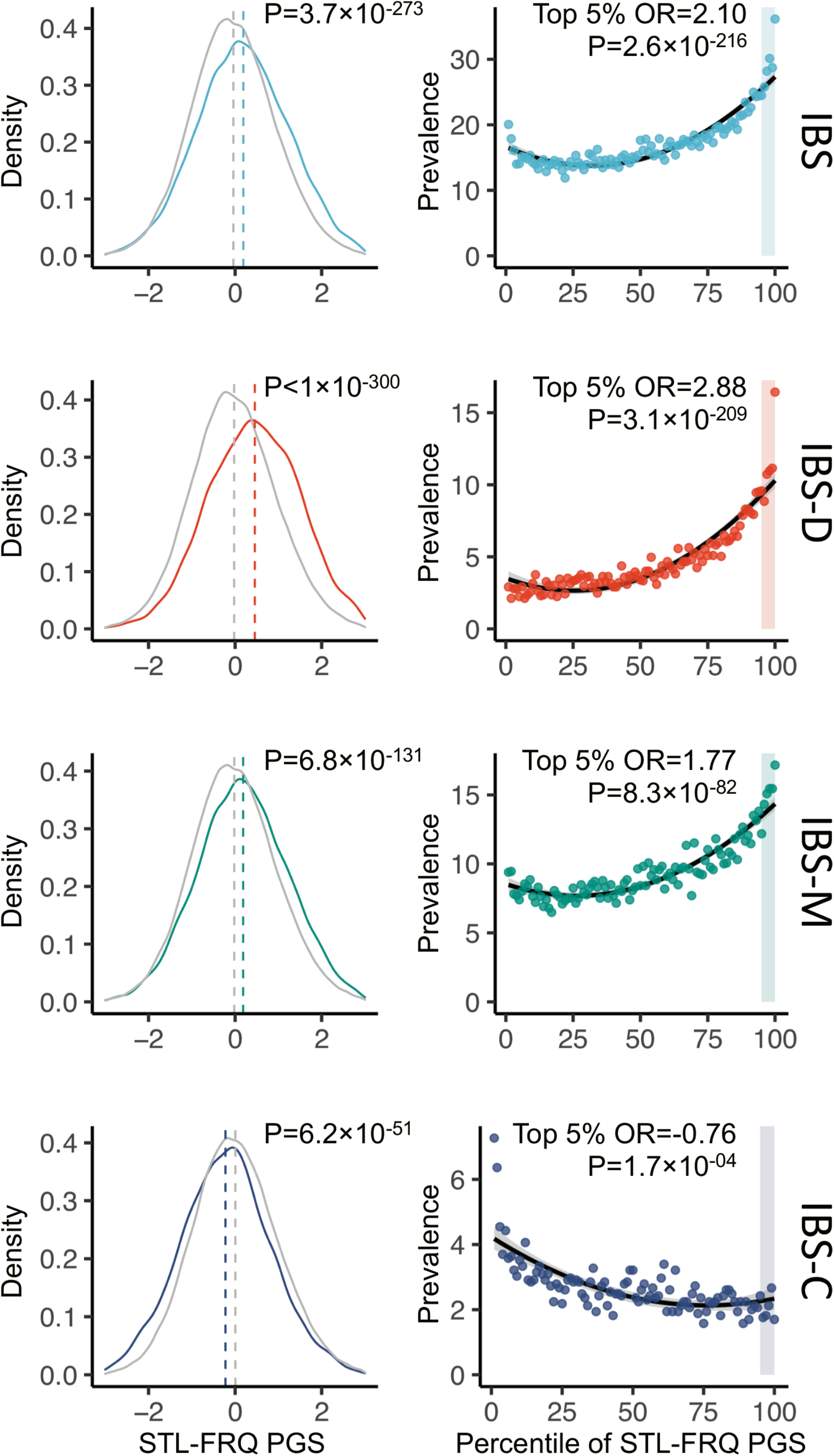
STL-FRQ polygenic scores (PGS) and IBS in UK Biobank. Results are reported (including statistical significance) in relation to PGS distribution in IBS cases vs controls (left panels, P-values from t-test), and in relation to the prevalence of IBS across PGS percentiles in the entire cohort (right panels; with top 5% of the distribution highlighted with shaded area, P-values vs the rest of the cohort from logistic regression). IBS and subtypes defined according to Rome III Criteria based on DHQ questionnaire data (see Methods).

IBS-D prevalence increased from 2.9% in the lowest to 16.4% in the highest PGS percentile, with individuals showing markedly increased IBS-D risk towards the tail of PGS distribution (ORs 4.14; P=1.2×10^−97^ for the top 1% and ORs 2.88; P=3.1×10^−209^ for the top 5% of the distribution, respectively; Figure 5 and Supplementary Table 8). However, although focused on a different trait (IBS), these analyses were performed on UK Biobank participants also included in the STL-FRQ GWAS (97% overlap), hence we further tested STL-FRQ PGS in the independent remainder of UK Biobank (N=291,496), in relation to combined IBS diagnoses available from touchscreen questionnaire (self-reported) and electronic medical records (ICD10 codes) (see Supplementary Methods). Although the prevalence of IBS defined by this approach was much lower (3.3%), and the diarrhea or other IBS subtypes could not be tested, similar results were obtained, thus replicating Rome-III findings: PGS values were significantly higher in cases than controls (respective means 0.042 and -0.001; P=2.9×10^−5^), IBS prevalence increased across PGS percentiles (3.6-4.2% bottom-top percentile range) and the risk of IBS was highest in the top 1% of the PGS distribution (1.29 OR; P=6.7×10^−3^; Supplementary Table 8).

## DISCUSSION

We report the results of a powered STL-FRQ GWAS, based on the meta-analysis of genetic and health-related data from five population-based cohorts. We undertook this study because of the known correlation between stool frequency and gut motility, whereas the latter cannot be feasibly studied in humans in numbers large enough for meaningful genetic investigations. Our approach therefore aimed at revealing relevant physiological pathways and mechanisms via indirectly measuring GI function based on suitable questionnaire data on bowel habits. A similar strategy was adopted in a previous study, however no significant results were obtained, likely due to the small size of the cohorts analyzed (total N=1281).[27] Studying almost 170,000 individuals, here we identify thousands of associations from 13 independent genome-wide significant loci, which harbor genes and DNA variants implicating pathways, cell types and mechanisms plausibly affecting human gut motility in health and disease.

Functional annotation and GWAS-downstream analyses suggest that genes from STL-FRQ loci are largely involved in neuropeptide and neurotransmitter signaling, sensory perception and control of motor function in the gut, which provides further evidence of the validity of our approach. These pathways are notoriously central to the ENS and its effects on GI motility, enabling bidirectional communication along the gut-brain axis.[28] Neuropeptides and neurotransmitters regulate gut behavior by propagating neuronal signals to the mucosal, immune and muscle systems, with excitatory and inhibitory effects on muscle contraction and peristalsis, among other functions. Our gene expression and cell-type enrichment analyses provide additional insight by harnessing the power of single-cell transcriptomics: exploiting RAISIN data from human colonic muscularis propria, we reveal how STL-FRQ genes are strongly enriched for their expression in enteric neurons, a specific pattern otherwise undetected at the whole tissue level. In particular, the enrichment appears to be more pronounced in putative excitatory and inhibitory motor neurons that have been associated with peristalsis and mechanosensation of gut distention (PEMN and PIMN subtypes expressing the mechanosensitive ion channel *PIEZO2*).[20] Further investigation of such expression patterns, coupled with functional characterization of STL-FRQ genes in specialized cell types and neuronal subtypes, may therefore contribute important insight into the exact mechanisms underlying neurogenic motor control in the gut, including dissecting specific classes of ENS neurons into their respective functional roles. This may aid the development of future therapeutic strategies to modify GI function and motility.

Individual genes most likely candidate to play an important role in the control of STL-FRQ also clearly point to the involvement of neuropeptide/neurotransimitter signaling pathways. This is best exemplified by the strongest association we detected in our GWAS meta-analysis at the *BDNF* locus on chromosome 13, which also replicated in CTT analyses of IBS patients. The association signal is mapped with relatively high confidence (>50% probability) to the rs12273363 marker, which is linked to multiple functional effects on *BDNF* expression: it has eQTL effects on an antisense transcript (*BDNF-AS*) that induces *BDNF* mRNA degradation,[25] and lies in a regulatory region previously shown to impart allele-specific, direct repression of *BDNF* promoter activity (with rs12273363 T a less active repressor).[29] *BDNF* is a neurotrophin expressed in the central and peripheral nervous systems, with neurotransmitter modulatory properties and a crucial role in neuronal growth, differentiation, survival and plasticity.[30] It has also been implicated in several diseases including major depression, bipolar disorder and other psychiatric conditions.[31] *BDNF* is recognized to influence many important gut functions, including sensation, motility, epithelial barrier, neuroprotection, and neuroplasticity.[32] Multiple lines of evidence indicate *BDNF* has prokinetic effects on gut motility, as shown by impaired peristalsis and delayed GI transit in *BDNF*+/- mice,[33] increased colonic myoelectric activity in *BDNF*-treated rats,[34] reduced *BDNF* colonic levels in patients with slow-transit constipation[35] and, notably, accelerated GI and colonic transit in individuals administered recombinant *BDNF* (r-metHuBDNF).[36] Hence, our findings are in line with these observations, in that the rs12273363 T allele associated with more frequent stools and shorter CTT has also been shown to induce stronger *BDNF* expression (weaker repressor).[29] Altogether, this suggests a *bona fide* role for *BDNF* in the genetically-determined modulation of human gut motility, and warrants new analyses of recombinant *BDNF* trials based on genotype stratification.

Our results also point to interesting candidate genes from other STL-FRQ loci where the association signal has been refined: *ACHE, FAXDC2* and *CDK18* all show eQTLs association with individual variants that have been fine mapped with >50% probability (respectively, rs4556017 on chromosome 7, rs13162291 on chromosome 5 and rs11240503 on chromosome 1). *ACHE* codes for an enzyme that hydrolyzes the neurotransmitter acetylcholine at neuromuscular junctions and is overexpressed in Hirschsprung’s disease,[37] while *FAXDC2* is a hydroxylase of fatty acids whose luminal concentrations are known to affect gut motility;[38,39] they are both expressed in enteric and motor neurons and therefore represent ideal functional candidates. *CDK18* encodes a protein kinase expressed in colonic M cells and BEST4+ enterocytes specialized in electrolyte and pH sensing,[20,40] hence its associated colon-specific eQTL may be relevant to colonic osmolarity and, consequently, transit.

Finally, strong functional candidates with a well-known role in GI motility map to additional loci where the association could not be attributed to specific variants. These involve additional neuropeptides/neurotransmitter systems, including alpha and beta calcitonin-gene related peptides (*CALCA* and *CALCB* genes) from the rs6486216 locus,[41] and the corticotropin-releasing hormone receptor (*CRHR1*) from the rs2732706 locus.[42] Altogether, these and previous observations made for STL-FRQ candidate causative genes are particularly interesting, in that they may provide rationale for future translational opportunities in the dysmotility syndromes. Several STL-FRQ associations were previously detected also in other health and disease-related traits, as from our cross-trait approach to interrogating publicly available GWAS data. Ten out of thirteen GWAS loci were already linked to lifestyle, anthropometric and disease-related traits (psychiatric conditions in particular). Broader evidence of genetic overlap with these conditions came from our LDSC analyses, which further highlighted shared genetic architecture with gastrointestinal diseases and often co-morbid neuroaffective traits,[43] among others. This likely reflects the recognized importance of the gut-brain axis, and suggests our results may be exploited to gain disease insight in addition to their relevance to better understanding the physiology of human gut motility. We explored this in relation to IBS, the most common FGID and the archetype of dysmotility syndromes, which also showed strongest correlation with STL-FRQ among all traits tested in the LDSC analysis.

Polygenic scores (PGS; calculated by summing multiple alleles weighted by their effect sizes, usually derived from GWAS studies), are an attractive way to capture an individual’s predisposition to develop a specific trait or disease, and hold strong potential for clinical translation and patient stratification. We computed PGS based on our STL-FRQ GWAS meta-analysis, and tested them in relation to IBS in the large UK Biobank cohort. STL-FRQ PGS were significantly higher in IBS cases vs asymptomatic controls defined according to Rome III criteria available for approximately 165,000 individuals, as well as in individuals with a doctor’s diagnosis of IBS (self-reported or in their medical records) compared to all other participants in the remainder of UK Biobank (almost 300,000 people). Individuals from the upper tail of the PGS distribution were more likely affected by IBS, and exposed to up to >4x higher risk of IBS-D compared to the rest of the population (in the top 1% of the distribution). Of note, at least in UK Biobank, the heritability of STL-FRQ (h^2^_SNP_ =0.073) appears to be higher than that of IBS (h^2^_SNP_ =0.037 on the liability scale, based on previous GWAS data on self-reported IBS).[15] This suggests that, once refined and further validated in independent cohorts, PGS derived from the simple STL-FRQ trait may ultimately contribute to an early identification, and eventual preventive treatment, of individuals at higher risk of developing IBS and other complex dysmotility syndromes.

Finally, our study has a number of limitations: i) stool frequency defined according to questionnaire data only equates to human gut motility to a certain extent, as its correlation with GI transit time has been shown to be weaker than, for instance, stool consistency; ii) current analyses could not take into account likely contributing environmental factors like diet, medications and others (whose related information was unavailable in most datasets); iii) relevant cell types and neuronal species have been identified and further classified here based on gene expression data, hence functional characterizations may be necessary to confirm specific mechanisms involved in the control of STL-FRQ and motility, as proposed; and finally iv) most STL-FRQ loci still require conclusive identification of the individual causative gene and variant(s). These issues can be addressed in future studies, and should therefore stimulate further investigation as follow-up to the novel findings reported here.

In conclusion, we identify loci harboring prioritized genes with a plausible role in GI motility, possibly acting via neurotransmission and similar pathways in specialized enteric neurons. The demonstrated relevance of these findings to IBS warrants further study for the identification of actionable pathomechanisms in the dysmotility syndromes.

## Data Availability

Full GWAS summary statistics will be uploaded to GWAS catalog online

## ACKNOWLEDGEMENTS

This research has been conducted using the UK Biobank Resource under Application Number 17435.

## AUTHOR CONTRIBUTORSHIP

MD and AZ: study concept and design; AA, LA, SW, GA, MP, DAH, NT, JR, AF, NAK, AR, AZ, MS, MC, MD: cohorts, patients characterization, data collection; FB, XL, CS, AP, AK, RB, TZ, HN, KGE: statistical analyses; FB, XL, CS, AP, AK, RB, LB, CE, LJ, MP, NT, JR, AF, NAK, AR, AZ, MS, MC, MD: data analysis and interpretation; MD: obtained funding, administrative and technical support, study supervision; FB and MD: drafted the manuscript, with input and critical revision from all other authors.

